# Autoantibodies Detected in MIS-C Patients due to Administration of Intravenous Immunoglobulin

**DOI:** 10.1101/2021.11.03.21265769

**Authors:** Peter D. Burbelo, Riccardo Castagnoli, Chisato Shimizu, Ottavia M. Delmonte, Kerry Dobbs, Valentina Discepolo, Andrea Lo Vecchio, Alfredo Guarino, Francesco Licciardi, Ugo Ramenghi, Emma Rey, Maria Cecilia Vial, Gian Luigi Marseglia, Amelia Licari, Daniela Montagna, Camillo Rossi, Gina A. Montealegre Sanchez, Karyl Barron, Blake M. Warner, John A. Chiorini, Yazmin Espinosa, Loreani Noguera, Lesia Dropulic, Meng Truong, Dana Gerstbacher, Sayonara Mató, John Kanegaye, Adriana H. Tremoulet, Pediatric Emergency Medicine Kawasaki Group, Eli M. Eisenstein, Helen C. Su, Luisa Imberti, Maria Cecilia Poli, Jane C. Burns, Luigi D. Notarangelo, Jeffrey I. Cohen

## Abstract

The autoantibody profile associated with known autoimmune diseases in patients with COVID-19 or multisystem inflammatory syndrome in children (MIS-C) remains poorly defined. Here we show that adults with COVID-19 had a moderate prevalence of autoantibodies against the lung antigen KCNRG, and SLE-associated Smith autoantigen. Children with COVID-19 rarely had autoantibodies; one of 59 children had GAD65 autoantibodies associated with acute insulin-dependent diabetes. While autoantibodies associated with SLE/Sjögren’s syndrome (Ro52, Ro60, and La) and/or autoimmune gastritis (gastric ATPase) were detected in 74% (40/54) of MIS-C patients, further analysis of these patients and of children with Kawasaki disease (KD), showed that the administration of intravenous immunoglobulin (IVIG) was largely responsible for detection of these autoantibodies in both groups of patients. Monitoring *in vivo* decay of the autoantibodies in MIS-C children showed that the IVIG-derived Ro52, Ro60, and La autoantibodies declined to undetectable levels by 45-60 days, but gastric ATPase autoantibodies declined more slowly requiring >100 days until undetectable. Together these findings demonstrate that administration of high-dose IVIG is responsible for the detection of several autoantibodies in MIS-C and KD. Further studies are needed to investigate autoantibody production in MIS-C patients, independently from IVIG administration.

## Introduction

Coronavirus disease 2019 (COVID-19) caused by the SARS-CoV-2 virus, is associated with a high rate of respiratory-related mortality [1]. Besides pulmonary disease, SARS-CoV-2 infection can result in acute injury to the kidney, liver, and heart, disseminated intravascular coagulation, rhabdomyolysis, and chronic fatigue [2]. The variability in clinical symptoms with COVID-19 likely involves sites of virus and viral antigen localization, vascular complications associated with the infection, the level of inflammation, and host factors including genetics, age, gender, and coexistence of other comorbid conditions [3]. In addition, the host immune response has a significant impact on disease severity and outcome [4, 5]. One major immune pathway involved in limiting SARS-CoV-2 infection is the type I interferon (IFN) pathway. Evidence for its role is based on genetic studies showing that individuals with inborn errors of type I IFN are at higher risk of severe COVID-19 [6]. Furthermore, neutralizing autoantibodies to IFN-α and/or -ω occur in approximately 10% of patients with severe COVID-19 pneumonia, especially men, and blunt effective clearance of the virus, likely contributing to death [7].

Compared to adults, most children and young adolescents with acute SARS-CoV-2 infection remain asymptomatic or have mild symptoms [8, 9]. However, a rare, severe condition, multisystem inflammatory syndrome in children (MIS-C), presents with high-grade fever, rash, multi-organ dysfunction, and elevated markers of inflammation, which typically occur 4-6 weeks after SARS-CoV-2 infection [10, 11]. MIS-C clinical manifestations resemble in part those of Kawasaki Disease (KD), a vasculitis typically affecting children. Children with MIS-C more frequently show myocardial involvement and gastrointestinal symptoms. The mechanisms involved in the pathogenesis of MIS-C are uncertain. Common treatments for MIS-C and KD include high dose intravenous immunoglobulin (IVIG), corticosteroids, and biologic agents [12]. In particular, IVIG, containing pooled IgG purified from >10,000 donors [13], is the standard first-line treatment for KD and is often administered to MIS-C patients.

Increasing efforts are being directed at understanding the differences in symptoms and complications seen in COVID-19 patients with the goal of improving treatment. One strategy is to identify biomarkers associated with, and predictive of, severe COVID-19 outcomes and involves analysis of soluble molecules in blood such as cytokine, chemokines, and other inflammatory markers [14-16]. From these and additional studies, a large body of evidence implicates the importance of the immune system in both controlling infection and promoting tissue damage. Several recent studies have identified autoantibodies in cross-sectional serum samples of adults and children with COVID-19 and children with MIS-C [17-23]. However, the possible confounding role of high-dose IVIG administration in the detection of such autoantibodies has not been carefully studied. Here, we assessed the presence of autoantibodies against a panel of autoantigens associated with known autoimmune diseases in adults and children with COVID-19. Our analysis showed a moderate prevalence of autoantibodies targeting the lung antigen KCNRG in adults, but not in children with COVID-19. We also demonstrate a high prevalence of autoantibodies associated with systemic lupus erythematosus (SLE)/Sjögren’s syndrome and autoimmune gastritis in the serum or plasma of patients with MIS-C, which we found to be associated with previous administration of high-dose IVIG and to persist for 40 to 120 days before they are cleared from the blood.

## Methods

### Patient populations

Deidentified plasma or serum samples were obtained from patients with COVID-19 or control subjects from multiple sites under an NIH IRB exemption after local IRB approval: Brescia, Italy Comitato Etico Provinciale (NP 4000 and NP 4408); Pavia, Italy (Comitato Etico Pavia Prot. 20200037677); Turin, Italy (Comitato Etico Interaziendale A.O.U. Città della Salute e della Scienza di Torino, Protocol: 00282/2020); Naples, Italy (Ethics Committee of the University of Naples Federico II, Protocol: 158/20); Chile (Comité Ético Científico Facultad de Medicina Clínica Alemana Universidad del Desarrollo, Santiago, Chile, Protocol: 2020-41); and Israel (Hadassah Medical Organization Institutional Review Board (IRB) for studies involving human subjects, Protocol: HMO-235-20). Additional MIS-C patients were enrolled from other sources including Stanford, California and Portland, Oregon, after written informed consent to NIH IRB-approved research protocols (NIH Protocols l1-1-0109, 18-I-0041 and 18-I-0128). Sera from children with KD or acute febrile illnesses were obtained at the University of California, San Diego under IRB approved protocols (UCSD #140220). All patients and parents gave written informed consent and assent as appropriate.

Serum samples from adults with COVID-19 (n=80) in Brescia, Italy were obtained from a higher number of male than female (48:32) patients with a median age of 55 years (interquartile range (IQR): 45-66), and a median time between blood draw and onset of symptoms of 25 days (IQR: 13-52). These adults had varying degrees of severity of disease (critical, severe, moderate, and mild) based on a previously described clinical rating system [24]. Serum samples from SARS-CoV-2 uninfected, healthy children (n=16) were obtained from Pavia, Italy (**Table 1**).

**Table 1.** Subject characteristics of the pediatric cohort (n=228)

|  | **N** | **Sex**<br>**(M:F)** | **Age**<br>**(years)<sup>a</sup>** | **SARS-CoV-2**<br>**PCR and/or**<br>**serology positive** | **Time from symptoms**<br>**onset to blood draw**<br>**(days)<sup>a</sup>** |
| --- | --- | --- | --- | --- | --- |
| **Healthy pediatric controls** | 16 | 7:9 | 6.5 [3-13] | NA | NA |
| **Acute febrile infection, unrelated to SARS-CoV-2 or Kawasaki Disease** | 15 | 4:11 | 10 [8-12] | NA | 8 [ 7-10] |
| **Kawasaki Disease-acute phase** | 39<sup>b</sup> | 29:10 | 9 [8-10] | NA | 8 [7-11] |
| **Kawasaki Disease-subacute phase** | 37<sup>b</sup> | 26:11 | 8 [7-11] | NA | 40 [37-45] |
| **Children with COVID-19** | 59 | 41:18 | 1 [0.7-9] | 59 (100%) | 2 [0-6] |
| **Children recovered from COVID-19** | 8 | 7:1 | 14 [11-15] | 8/8 | 52 [30-142] |
| **MIS-C** | 54 | 29:25 | 6 [2-11] | 54 (100%) | 3 [1-9]<sup>c</sup> |

<sup>a</sup> Values in square brackets represent the median and interquartile range.
<sup>b</sup> Ten of the same patients with Kawasaki disease in the acute and subacute phase were represented.
<sup>c</sup> For MIS-C, time is defined as the day of the first blood draw obtained after hospitalization.
NA- Not applicable

Serum/plasma samples from children approximately 8 days after onset of acute febrile infections unrelated to KD or SARS-CoV-2 (n=16), from children during the acute phase of KD (median time of 9 days after disease onset, n=39), and from children during the subacute phase of KD (median time of 40 days after disease onset, n=37) were obtained from the University of California, San Diego. Samples from children with COVID-19 and MIS-C were from multiple locations in the US, Israel, Italy, and Chile. Among the children with COVID-19 without MIS-C, 59 were in the acute phase of COVID-19 and 8 had recovered from COVID-19 that had occurred approximately two months earlier. All patients with MIS-C (n=54) were SARS-CoV-2 PCR and/or antibody positive (**Table 1**). Forty-four percent of the patients with MIS-C had 2 or more longitudinal samples that were obtained from the time of initial hospitalization to several weeks later.

### LIPS antibody and autoantibody testing

Luciferase Immunoprecipitation Systems (LIPS) assays use a 96-well format to detect antibodies to both conformational and linear epitopes in a fluid phase system [25]. A key aspect of the LIPS technology is the use of custom luciferase-antigen fusion proteins, containing the antigen of interest, which provide high sensitivity and specificity and a wide dynamic range of detection. COVID-19 subjects were evaluated by LIPS for serum/plasma antibodies against the nucleocapsid and spike proteins of SARS-CoV-2 as previously described [26]. Panels of autoantigen targets used for LIPS testing have been previously described and validated in other published and unpublished autoimmune cohorts, including SLE [27], Sjögren’s syndrome [28], systemic sclerosis [29], myositis [30], ANCA-associated vasculitis, autoimmune polyendocrinopathy-candidiasis-ectodermal dystrophy (APECED) [31], type 1 diabetes [32, 33], autoimmune encephalitis, and membranous nephropathy [34]. A selected panel of autoantigens representing both ubiquitous and tissue-specific antigens were first tested in adults and included: Ro52, Ro60, La, RNP-A, Sm-D3, Jo-1, gastric ATPase (ATPB4 subunit), LGI1, U170K, CENP-A, RNAP-K, BPIFB1, defensin 5A, KCNRG, glutamic acid decarboxylase (GAD65), gastric inhibitory factor (GIF), and M-type phospholipase A_2_ receptor (PLA2R). Based on the results of the adult COVID-19 study and the limited availability of serum for the COVID-19 children with and without MIS-C, a smaller number of autoantigens were selected for study in these children. Due to a single case showing GAD65 autoantibodies in a patient with symptoms of acute onset of type 1 diabetes, follow-up LIPS studies were performed to determine immunoreactivity to IA2 and ZNT-8, two autoantigens associated with diabetes mellitus. While healthy adult blood donor samples were used for to determine negative cut-off values for LIPS assays in adult COVID-19 patients, the control children without COVID-19 were used to determine the cut-off values in children with KD, COVID-19 and MIS-C. The value derived from the mean plus three standard deviations of the controls were used to set the cut-off for determining seropositivity [28, 32]. For the gastric ATPase, one elevated outlier from the control group of children was excluded in determining the cut-off value.

LIPS testing was also used to evaluate the autoantibody levels in three different batches of IVIG. For these experiments, the stock 10% IVIG solution (Gamunex®, Grifols Therapeutics Inc.) was diluted 1:10 in PBS to achieve approximately the same concentration of IgG present in human sera (20 μg/μL). In addition to testing the IVIG preparation for autoantibodies against Ro52, Ro60, La and the gastric ATPase, several additional autoantigens were tested including Jo-1 [30] and IL-1 alpha [35].

### Statistical analysis

GraphPad Prism software (San Diego, CA) was used for analyzing the antibody levels in this study. Median antibody levels, expressed as mean log_10_ light units (LU) and interquartile range (IQR), were calculated and presented as antilog values. The non-parametric Mann-Whitney *U* statistical test was used for comparison of antibody levels in the control and COVID-19 patient groups. The Fisher’s exact test was used to evaluate the statistical significance of the prevalence of autoantibodies in patient groups and controls. JMP® 14.0.0 was used to illustrate the kinetics of antibody levels over time using a smoothing spline (cubic spline) with a λ of 0.77 and a shaded area representing the bootstrap confidence region for each fit.

## Results

### Characteristics of adults and children with COVID-19 and children with MIS-C

Plasma from adults and children with acute COVID-19 and children with MIS-C were obtained from multiple geographical sites including the US, Italy, Israel, and Chile. The median age of the MIS-C children (n=54) was 6 years (IQR; 2 to 11 years) which was older than in children with COVID-19 (median age 1 year, IQR; 0.7 to 9 years) **(Table 1)**. Furthermore, there was an approximately equal number of males and females in MIS-C patients, while the children with COVID-19 had a male predominance (69%).

Sixty-one percent of the patients with MIS-C were Hispanic/Latino, 32% were Caucasian, and 7% were black (**Table 2**). The most common symptom in the MIS-C cohort was fever (100%) followed by gastrointestinal symptoms of abdominal pain, vomiting, and diarrhea (91%). Of the total cohort of MIS-C patients, 87% were treated with corticosteroids and 70% were given IVIG.

**Table 2.** Demographics and clinical characteristics of the MIS-C cohort (N=54)

|  |  |
| --- | --- |
| **Sex (M:F)** | 29:25 |
| **Age<sup>a</sup>** | 6 [2-11] |
| **Ethnicity** |  |
| Hispanic/Latino | 33 (61%) |
| Caucasian | 17 (32%) |
| Black/African American | 4 (7%) |
| **Clinical manifestations** |  |
| Fever | 54 (100%) |
| GI symptoms | 49 (91%) |
| Myocarditis/heart failure | 37 (68%) |
| Coronary artery involvement | 10 (18%) |
| Shock (distributive/cardiogenic) | 33 (61%) |
| Neurologic involvement | 19 (35%) |
| Respiratory Symptoms | 19 (35%) |
| Rash/cutaneous manifestations | 33 (61%) |
| **Laboratory values at the admission** |  |
| ALC <1.5 x10<sup>9</sup> cells/L | 30/52 (58%) |
| PLT <150 x10<sup>9</sup> cells/L | 17/52 (32%) |
| CRP >10 mg/L | 49/50 (98%) |
| ALT >40 U/L | 24/48 (50%) |
| Ferritin >500 µg/L | 16/34 (47%) |
| D-dimer >500 µg/L | 40/50 (80%) |
| **Highest level of care and outcome** |  |
| Hospitalization | 54 (100 %) |
| ICU Admission | 22 (40%) |
| Death | 0 |
| **Treatments** |  |
| Corticosteroids | 47 (87%) |
| IVIG | 38 (70%) |
| Corticosteroids AND IVIG | 34 (62%) |
| Biologics | 4 (7%) |
| Inotropes | 20 (37%) |

<sup>a</sup>Numbers represent median values (unless otherwise specified) and numbers in square parentheses represent 1st and 3rd quartiles.
ALC, absolute lymphocyte count; ALT, alanine aminotransferase; CRP, C-reactive protein; GI, gastrointestinal; ICU, intensive care unit; IVIG, intravenous immunoglobulin; PLT, platelets.

### Autoantibodies in adults with COVID-19

A cohort of adults with COVID-19 and with different levels of clinical severity was analyzed for autoantibodies against known antigens associated with multiple autoimmune diseases. A small number of COVID-19 patients had antibodies against Ro52, Ro60, and La comprising the SSA and SSB rheumatological autoantigens (**Figure 1 A-C**). Only one subject had autoantibodies to all three targets and no correlation of these autoantibodies was seen with disease severity. Testing of two SLE-associated antigens showed a lack of autoantibodies against RNP-A (**Figure 1D)**; however, there was a high frequency of Sm-D3 autoantibodies in 34% (27/80) of adult COVID-19 patients (**Figure 1E)**. Approximately 60% of the severe patients had Sm-D3 autoantibodies compared to 4%, 43%, and 27% in the subjects with critical, moderate, and mild disease, respectively (**Figure 1E)**. The high frequency of autoantibodies in severe and moderate disease compared to critical was statistically significant (p<0.004). Low levels of autoantibodies to KCNRG, a lung antigen, were found in 30% (24/80) of adult COVID-19 patients, but were significantly enriched in the patients with severe or moderate disease and generally absent from patients with critical or mild disease (*P*<0.0001) (**Figure 1F**). Autoantibodies against GAD65, an autoantigen associated with type 1 diabetes and autoimmune encephalitis, was also detected in 8% of the severe and 4% of the critical patients (**Figure 1G**). Finally, high levels of autoantibodies against gastric ATPase, which are associated with autoimmune gastritis, were detected in approximately 9% of the COVID-19 patients, but the presence of these autoantibodies did not correlate with clinical severity (**Figure 1H**). Testing of antibodies to several other autoantigens including CENP-A, PLA2R, and LGI1 that are found in patients with systemic sclerosis, membranous nephropathy, and limbic encephalitis, respectively, revealed little or no autoantibodies in any of COVID-19 adults (data not shown). Taken together, these results suggest that adults with COVID-19 had a diverse repertoire of serum autoantibodies to known autoantigens and at least two of them, the lung KCNRG and stomach gastric ATPase, might reflect infection-related damage to these tissues in selected patients.

**Figure 1.**
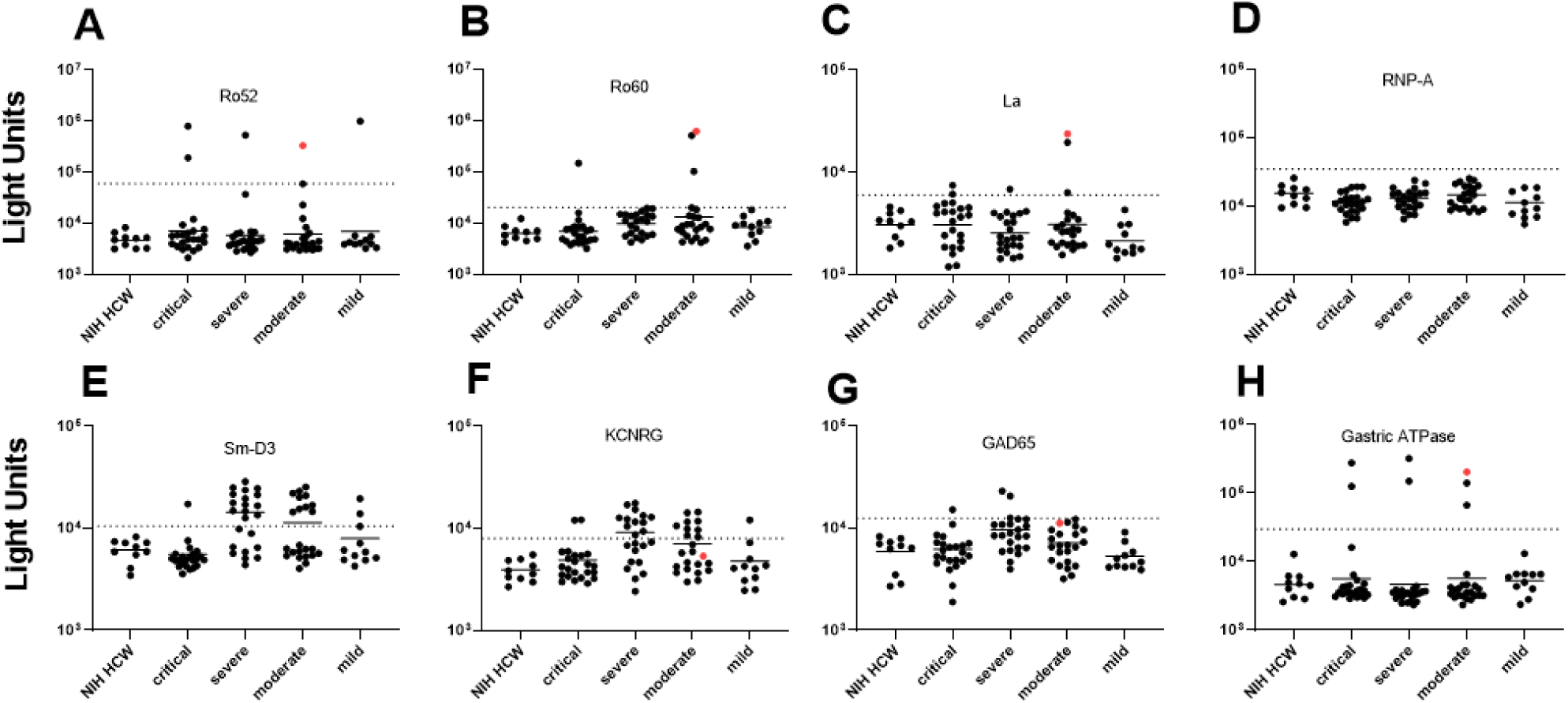
Autoantibodies against known autoantigens in adults with COVID-19 not receiving IVIG. Autoantibody levels against eight well known autoantigens were determined in adults with COVID-19 having varying levels of disease severity: critical, severe, moderate, and mild. Each symbol represents a sample from an individual control subject (SARS-CoV-2 uninfected health care worker (NIH HCW), a patient with COVID-19 (critical, severe, or moderate, or mild). Autoantibody levels are plotted in light units on a log_10_ scale. The dashed lines represent the cut-off level for determining positive antibody levels for each autoantigen as described in the Methods. Samples from a single patient with moderate disease severity showing antibodies to multiple autoantigens are shown by the red dots.

### Autoantibody profiles of children with COVID-19 or MIS-C

Levels of autoantibodies were also measured in cross-sectional and longitudinal serum samples of children with COVID-19 or with MIS-C, with many (70%) of the latter receiving IVIG. As controls, plasma samples collected from healthy children without SARS-CoV-2 infection were used as controls and to determine cut-off values for these autoantibodies. Approximately 50% of MIS-C children had Ro52, Ro60, and/or La autoantibodies (**Figure 2A-C**), which were nearly absent in children with COVID-19 (1/59) and in children (0/8) who had recovered from COVID-19. The one child with COVID-19 that was seropositive for autoantibodies had received IVIG for autoimmune thrombocytopenia six days before the serum sample was collected. In MIS-C, the high prevalence of autoantibodies against Ro52, Ro60, La and the gastric ATPase were often found together in the same individuals. The relative antibody levels were comparable to that seen in some patients with SLE or Sjögren’s syndrome [27, 28] and were on average over 10 times higher than the cut-off value. The prevalence of autoantibodies against Ro52, Ro60, and La antigens among the individual MIS-C patients was 57% (31/54), 67% (36/54), and 59% (32/54), respectively, and these autoantibodies were often present at more than one time point. None of the recovered COVID-19 children, and only one child at two different time points with COVID-19 had all three autoantibodies against Ro52, Ro60, La, and gastric ATPase autoantibodies. Autoantibodies against gastric ATPase were also present in 74% (40/54) of the children with MIS-C (**Fig 2D**). The median level of these gastric ATPase autoantibodies in MIS-C children was 122,705 LU (IQR 11,810-253,100) which was approximately 10-fold higher than in children with COVID-19 (8790 LU; IQR 7,567-10,670) and 12-fold higher than in uninfected children (9,825 LU; IQR 9190-11,314). Examination of whether autoantibody positivity against Ro52, Ro60, La, and gastric ATPase were associated with clinical features of the MIS-C patients revealed that patients with autoantibodies had more severe disease based on a statistically higher frequency of ICU admission than those without autoantibodies (*P*=0.0003) and all were treated with IVIG.

**Figure 2.**
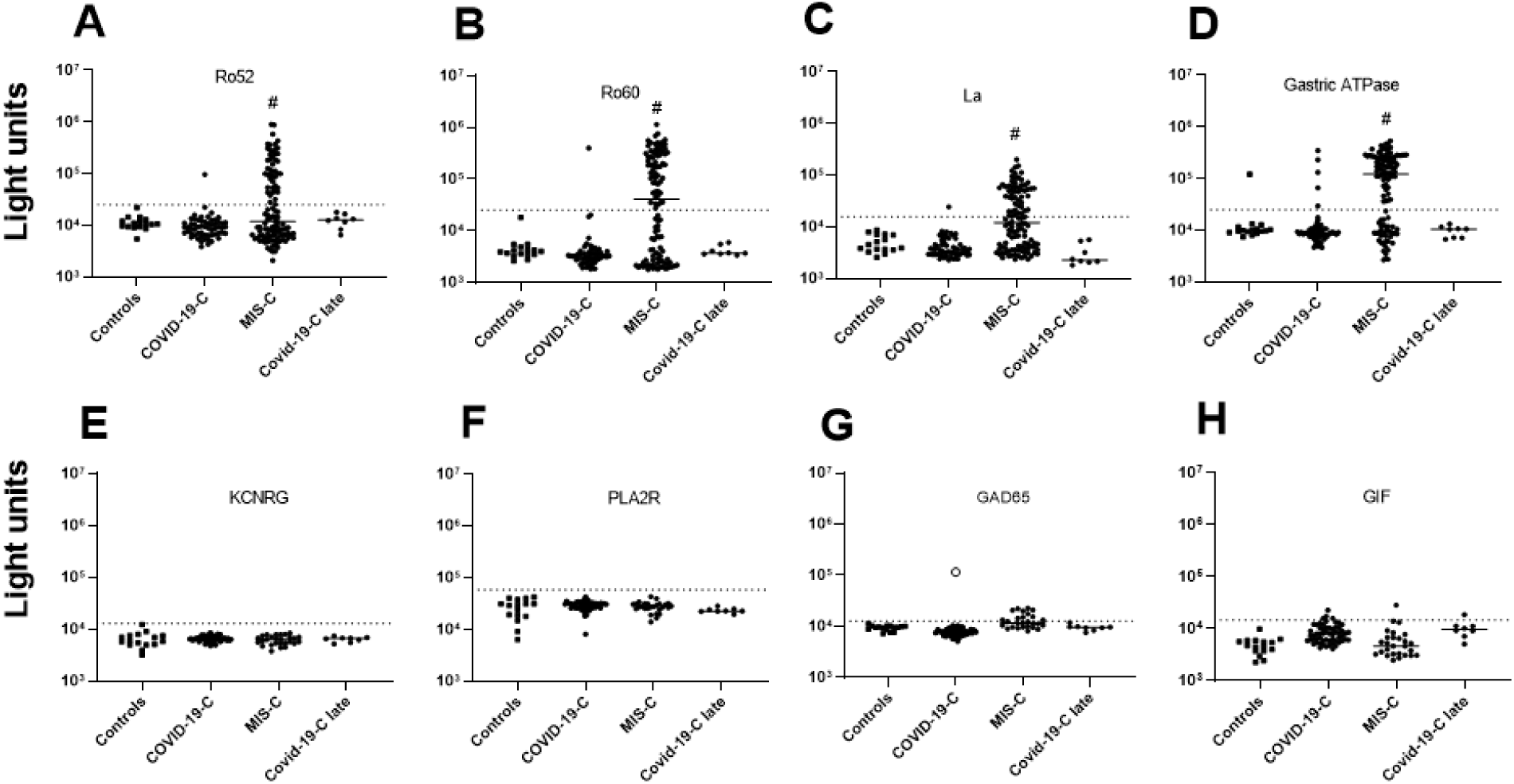
Many children with MIS-C treated with IVIG show a high prevalence of autoantibodies against Ro52, Ro60, La, and gastric ATPase. **(A-D)** Autoantibody levels against Ro52, Ro60, La, and gastric ATPase were determined in 16 children not infected with SARS-CoV-2 (Controls), 59 children with COVID-19 (COVID-19-C), 54 children with MIS-C, and 8 children who recovered from acute COVID-19 (COVID-19-C late). Each symbol represents a sample from an individual patient or different time points from an individual patient (**E-F)** Autoantibody levels against four autoantigens, KCNRG, PLA2R, GAD65, and GIF were tested in the same children as in panels A-D, except for MIS-C children where a single sample from the time of peak Ro52, Ro60, La, and/or gastric autoantibodies was used. Autoantibody levels are plotted on the Y-axis in light units on a log_10_ scale. The dashed lines represent the cutoff levels for determining positive autoantibody levels for each target antigen as described in the Methods. The sample from the child with acute COVID-19 with high levels of GAD65 autoantibodies is denoted by the open black circle. Statistically significant higher autoantibody levels seen in MIS-C patients treated with IVIG compared to the other groups are shown by the hash tag.

Samples from children with COVID-19 or MIS-C were also examined to determine if there was immunoreactivity against additional autoantigens including KCNRG, PLA2R, GAD65, and gastric intrinsic factor (GIF). No autoantibodies were detected above the cut-off values against the lung (KCNRG) and kidney (PLA2R) antigens (**Figure 2E and F**). High levels of GAD65 autoantibodies were detected in an adolescent girl with acute SARS-CoV-2 infection who had not received IVIG **(Figure 2G)**. Review of the medical records indicated that this child with COVID-19 was hospitalized with severe ketoacidosis and required intravenous insulin. Additional autoantibody testing against other type 1 diabetes-associated autoantigens including IA2, and Znt-8, was negative. These results suggest the possibility that acute COVID-19, at least in one child, may have contributed to an immune response triggering type 1 diabetes. Autoantibodies against GAD65 were also detected in approximately 26% of children with MIS-C, but their levels were only slightly above the cut-off threshold **(Figure 2G)**. Importantly, all the GAD65 seropositive MIS-C children had received IVIG prior to blood sampling. Testing for antibodies against GIF, which is produced by parietal cells and whose loss is associated with B12 deficiency, showed only sporadic low levels of GIF autoantibodies in children with COVID-19 and MIS-C (**Figure 2H**).

No autoantibodies against IA2 and defensin-5A (a self-antigen that is expressed in the ileum and is targeted by autoantibodies in patients with autoimmune polyendocrinopathy-candidiasis-ectodermal dystrophy patients) were detected in children with COVID-19 or MIS-C (data not shown).

### Ro52, Ro60, La, and gastric ATPase autoantibodies in MIS-C and KD are due to IVIG administration

Based on the high prevalence of Ro52, Ro60, La, and gastric ATPase autoantibodies in MIS-C, additional testing was performed to determine if these autoantibodies could help to distinguish MIS-C from KD and whether their presence could be affected by previous administration of high-dose IVIG. To address this, we analyzed serum/plasma from control children (n=8), children with acute febrile infections other than COVID-19 (n=15), children with MIS-C before they received IVIG (n=10), children with KD prior to receipt of IVIG (n=39), and children with KD after receiving IVIG (n=37). In addition, three batches of IVIG were tested for autoantibodies. Only one child with acute febrile illness not related to COVID-19, who was later diagnosed with SLE, showed Ro52, Ro60, and La autoantibodies, but not gastric ATPase autoantibodies (**Fig. 3**). Interestingly, no MIS-C children who had not received IVIG had autoantibodies to any of the four antigens (**Fig. 3**). Moreover, children with acute KD, who had not yet received IVIG, showed no autoantibodies against Ro52, but two showed seropositivity for gastric autoantibodies and a third patient had both Ro60 and La autoantibodies. In contrast, subacute KD patients from later time points, all of whom received IVIG in the previous 6 weeks, showed a very high rate of autoantibodies against Ro52, Ro60, La, and gastric ATPase with a frequency of 46%, 68%, 35% and 86%, respectively (**Fig. 3**). Five subacute KD patients showed no detectable autoantibodies. Testing of three different IVIG preparations diluted 1:10, which approximates the IgG antibody concentration in human serum showed remarkably high levels of autoantibodies against the four autoantigens. The median levels of Ro52, Ro60, La, and gastric ATPase autoantibodies approximated the highest levels seen in the KD patients with geometric mean levels of 125,000, 359,300, 120,000 and 117,800 LU, respectively (**Fig. 3**). In addition, testing of two commonly found autoantibody targets, IL-1α and Jo-1, that have been observed in both healthy persons and patients with myositis, respectively, revealed low level autoantibodies in IVIG just above the cut-off value derived from control children (data not shown). Together these results suggest that the Ro52, Ro60, La, and gastric ATPase autoantibodies observed in MIS-C and KD patients were due to prior infusion of high-dose IVIG and that other autoantibodies found in IVIG preparations may contribute to anomalous autoantibody seropositivity,

**Figure 3.**
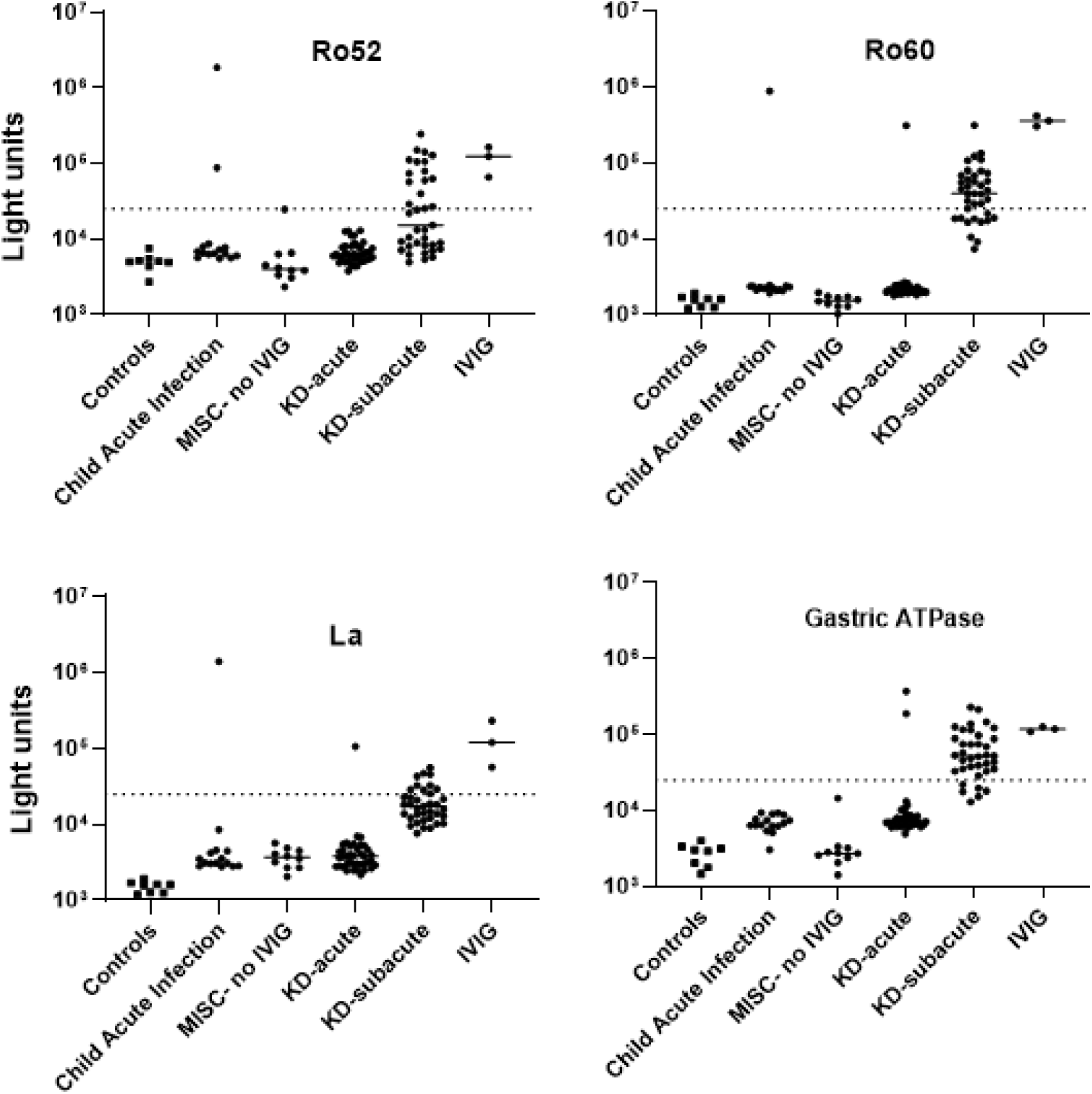
Autoantibodies in the serum of patients with MIS-C and Kawasaki disease (KD) are due to administration of IVIG. Autoantibody levels are plotted in light units on a log_10_ scale. The dashed lines represent the cut-off level for determining positive antibody titers as described in the Methods. Results from 8 sera from children not infected with SARS-CoV-2 (Controls), 15 sera from children with acute febrile infection not due to COVID-19 or KD (child acute infection), 10 sera from patients with MIS-C who did not receive IVIG (MISC-no IVIG), 39 sera from children with acute KD (KD-acute), 37 sera children with subacute KD (KD-subacute), and three preparations of IVIG are shown.

### Longitudinal analysis of SARS-CoV-2 autoantibodies in children with MIS-C and KD

To formally assess if the Ro52, Ro60, La, and gastric ATPase autoantibodies detected in the children with MIS-C and KD were due to IVIG, serial serum samples were tested from three MIS-C and three KD patients that were collected both before and after receiving IVIG. Analysis of the sera of MIS-C and KD patients before IVIG administration revealed that none of the patients had Ro52, Ro60, La, and gastric ATPase autoantibodies (**Figure 4**). However, serum samples taken within 24 hours after IVIG administration in these patients showed that they all contained high levels of autoantibodies. Furthermore, analysis of longitudinal samples from these six patients demonstrated high levels of these autoantibodies that persisted over the next 12 days (**Figure 4**). Taken together, these results confirm that the Ro52, Ro60, La, and gastric ATPase autoantibodies observed in these patients are directly due to administration of IVIG.

**Figure 4.**
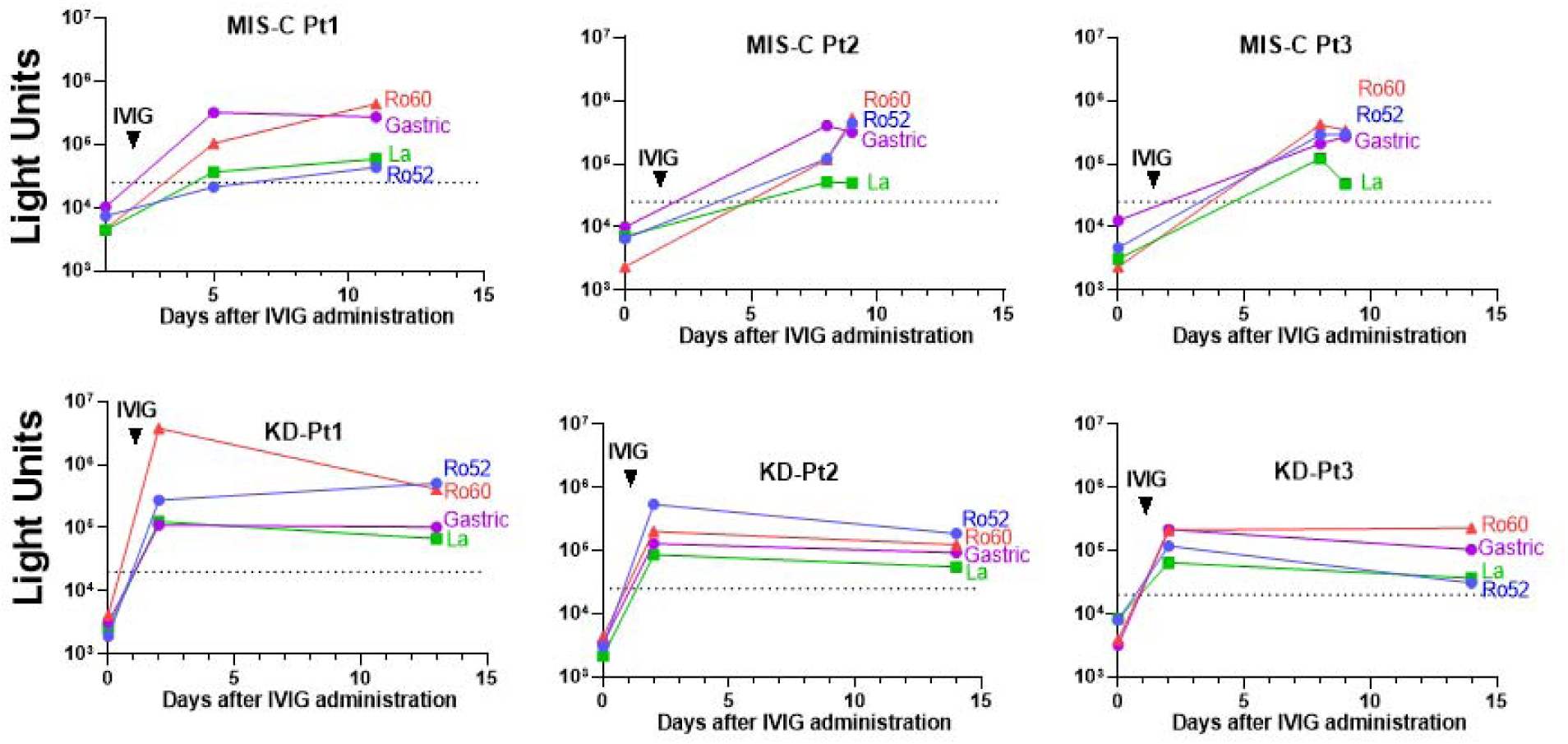
Longitudinal autoantibody profiles before and after administration of IVIG in MIS-C and KD patients. Antibody levels were determined in serial samples from representative MIS-C (MIS-C, Pt1-3) and KD patients (KD, Pt1-3), in which time zero represents the day of hospital admission. The autoantibody profiles for Ro52 (blue), Ro60 (red), La (green), and gastric ATPase (purple) are shown by the colored lines, in which the cut-off value for determining positivity is shown by the dotted lines. The arrow shows the day when IVIG was begun.

To accurately explore the kinetics of decay of autoantibodies, we assessed levels of Ro52, Ro60, La, and gastric ATPase autoantibodies in 21 MIS-C patients for which samples were available at two or more time points after IVIG administration (**Figure 5**). Based on the known time of IVIG administration, the Ro52, Ro60, and La autoantibodies showed a relatively rapid decline back to the seronegative cut-off value by 35-60 days. However, the gastric ATPase autoantibody decline trajectory was generally more delayed, in that most of the samples did not approach the seronegative cut-off value in the available serial samples. Modeling of gastric ATPase autoantibodies suggested a slower decay trajectory lasting >100 days.

**Figure 5.**
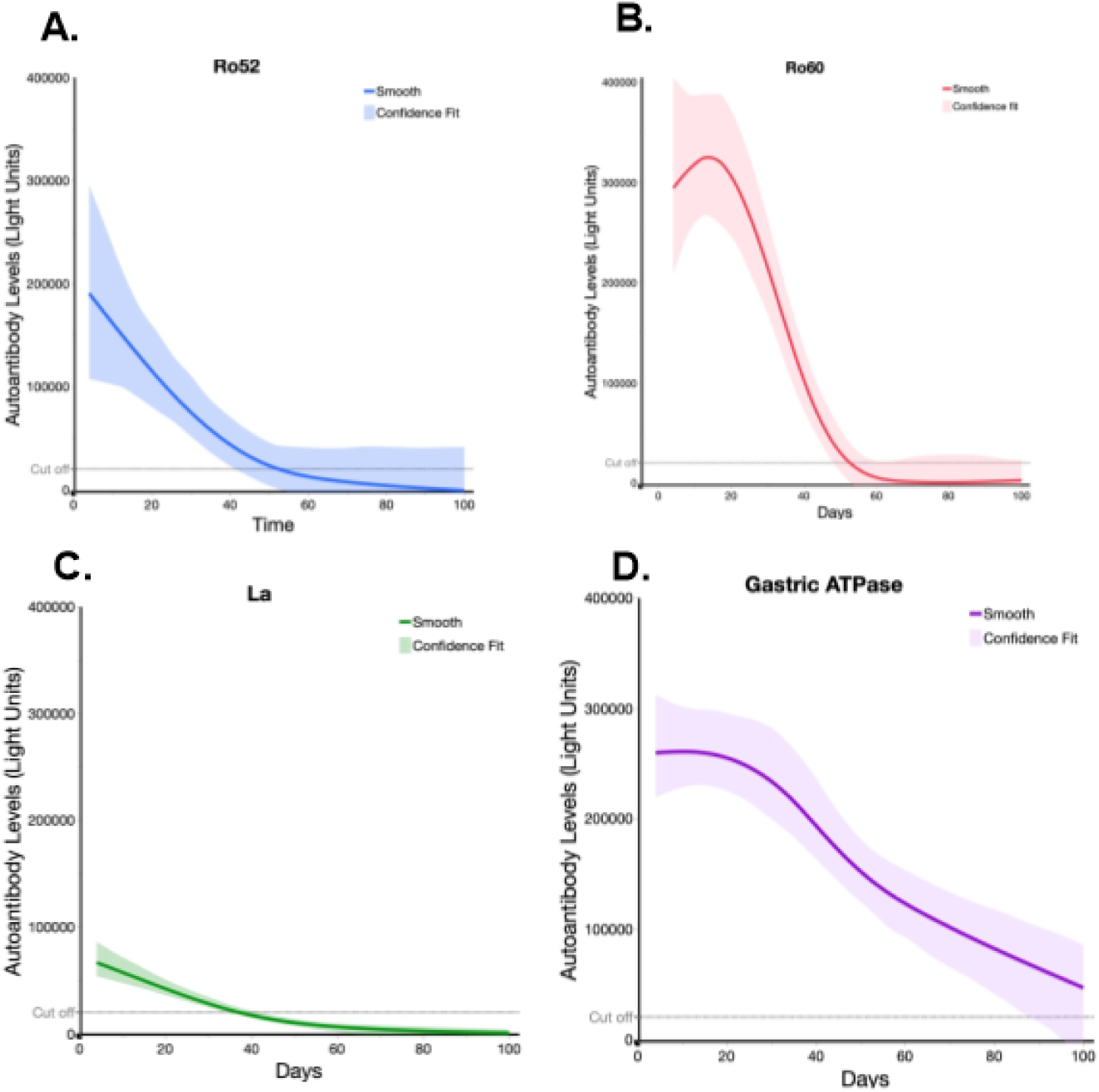
Ro52, Ro60, La, and gastric ATPase autoantibody decay in patients with MIS-C receiving IVIG. Decay plots were generated for Ro52, Ro60, La, and gastric ATPase autoantibody data from the MIS-C (n=21) patients having two or more different time points. Day 0 is the time the patients received IVIG. The solid-colored line for each autoantigen indicates the decay obtained using JMP® 14.0.0 and a smoothing spline (cubic spline) with a λ of 0.77 with the shaded area representing the bootstrap confidence region for each fit. The dotted lines indicate the cut-off value for determining seronegativity for each autoantigen.

## Discussion

Here we evaluated the prevalence of autoantibodies in adults and children with COVID-19 and children with MIS-C. Autoantibodies commonly found in autoimmune disease were chosen as target antigens because of their known association with different types of tissue damage. Analysis of adult patients with COVID-19 showed a moderate frequency of autoantibody positivity against several autoantigens including the lung protein KCNRG (30%) and the SLE antigen Sm-D3 (34%). Several other autoantigens, including Ro52, Ro60, La, and GAD65 were detected less frequently (< 8%) in the adults with COVID-19. While the levels of KCNRG autoantibodies seen in adults with COVID-19 occurred at a higher frequency than that seen in adults with ARDS (23%) or sepsis (25%) [36], they were lower than those reported in other diseases such as APECED complicated by pneumonitis (29%) [37]. Detection of autoantibodies to KCNRG is consistent with the lung injury and dysfunction that occur in many hospitalized COVID-19 patients. We found that some adults with COVID-19 had elevated antibodies to Sm-D3 as to well as Ro52, Ro60, and La. While RNP-A and Sm-D3 autoantibodies usually cluster together in patients with SLE [27, 38], only Sm-D3 autoantibodies were seen in adult COVID-19 patients. Moreover, both KCNRG and Sm-D3 autoantibodies were enriched in COVID-19 adult patients with severe and moderate disease, but not in critically ill COVID-19 patients. While the presence of specific autoantibodies may be driven by the release of the proteins that these antibodies target from damaged or dying cells in combination with immune activation and plasmablast expansion [39], possible explanations for the paradoxical lack of autoantibodies in critically ill patients is that these patients have a immunodeficiency state that can be similarly seen in late stages of sepsis or they might have received immunosuppressive medications that could have blunted their antibody response.

In contrast to adults, most children with COVID-19 demonstrated little or no autoantibodies against KCNRG, Sm-D3, and the other autoantigens we tested. The relative differences in the frequency of autoantibodies between children and adults is likely impacted by age-related differences in immunological function and increased comorbidity seen in adults [8]. One child with COVID-19 was found to have very high levels of GAD65 autoantibodies, a well-known marker of type 1 diabetes. The child presented with ketoacidosis and required insulin upon hospital admission. Although viruses have long been implicated in pancreatic beta cell destruction in type 1 diabetes [40, 41], recent studies show that SARS-CoV-2 can infect, replicate, and destroy beta cells that produce insulin in pancreatic islets [42, 43]. SARS-CoV-2 infection has been reported to induce both autoantibody-positive [44] and autoantibody-negative diabetes [43, 45]. SARS-CoV-2 infection has also been implicated in triggering the acute onset of several other autoimmune diseases, including encephalitis [46, 47], Guillain-Barre syndrome [48], and myasthenia gravis [49]. Based on these and other studies, our patient with SARS-CoV-2 associated type 1 diabetes further highlights that the virus may be a risk factor associated with the future onset of diabetes or other autoimmune diseases in children.

Children with MIS-C have inflammation involving multiple organ systems, which is usually not observed in children with COVID-19. KD typically present in in childhood and shares certain clinical features with MIS-C including fever, skin, mucous inflammation, and increased inflammatory markers [50]. One feature of MIS-C that is usually not seen in KD is the high frequency of gastrointestinal symptoms including abdominal pain, vomiting, and diarrhea [10, 11]. Several groups have reported the presence of a broad range of tissue-specific and non-organ-specific autoantibodies in MIS-C [17, 18, 22, 23]. Our initial autoantibody analysis suggested the possibility that there might be autoantibodies in MIS-C cases that were associated with various rheumatological diseases and autoimmune gastritis. However, further evaluation revealed that the autoantibodies against Ro52, Ro60, La, and gastric ATPase in the MIS-C patients’ sera were due to IVIG administration. Approximately 57% of the patients with MIS-C had all four autoantibodies, and the presence of these autoantibodies in MIS-C directly tracked with the administration of IVIG administration.

Multiple lines of evidence established that the autoantibodies against Ro52, Ro60, La, and gastric ATPase seen in MIS-C and KD children was due to IVIG administration. First, serum samples from both MIS-C and KD patients who did not receive IVIG did not have autoantibodies against Ro52, Ro60, La, and gastric ATPase. Among children with other acute febrile infections who did not receive IVIG, only one child had these autoantibodies, a child who was later diagnosed with SLE. In contrast, known KD and MIS-C children who received IVIG, showed a very high frequency (86-100%) of autoantibodies to these autoantigens. The few seronegative cases were likely confounded by different batches of IVIG and collection of serum samples weeks after IVIG administration. Second, our studies with multiple serial samples from both MIS-C and KD before and after IVIG revealed that the autoantibodies were only found after IVIG administration. Third, testing of three different lots of IVIG revealed high levels of autoantibodies against Ro52, Ro60, La and gastric ATPase, although the relative levels of these autoantibodies varied from the different lots of IVIG. These findings of existing autoantibodies in IVIG are also consistent with other published findings reporting autoantibodies in IVIG or in patients who received IVIG [51-55]. For example, SSA antibody, comprising autoantibodies against Ro52 and Ro60, has previously been detected in IVIG preparations [51], and Ro52 was identified as the most abundant autoantibody present in IVIG preparations used to treat neurological patients [52]. In addition, autoantibodies associated with other autoimmune diseases have been detected in IVIG preparations given to patients, including autoantibodies against GAD65 [53], the acetylcholine receptor [55] and desmosomal proteins [54]. One explanation for the seropositive autoantibodies in IVIG against these Ro52, Ro60, and La relates to the large IVIG donor pool who harbor high levels of these autoantibodies because they likely include samples from asymptomatic or pre-symptomatic individuals with current or future onset of SLE, Sjögren’s syndrome and other autoimmune conditions. The high levels of gastric ATPase autoantibodies seen in IVIG are likely due to donors not only having autoimmune gastritis but with other autoimmune diseases including type I diabetes, Sjögren’s syndrome, and SLE in which these antibodies have been detected [56]. Our results and work of others [54] also suggests that commercial IVIG preparations may differ in autoantibody concentrations due to differences in donor pools and manufacturing. Taken together these results show that IVIG given to MIS-C patients contains a variety of autoantibodies and without testing before IVIG treatment or allowing sufficient time after the dose of IVIG to allow decay of these antibodies before repeat testing, can be mistakenly interpreted as associated with the manifestations of MIS-C. Based on our findings, further studies are needed to investigate the nature, prevalence, and possible significance of autoantibody production in MIS-C independently from IVIG administration.

Three published studies have used antigen arrays for autoantibody discovery in MIS-C [17, 18, 22]. It is worth noting that none of these three publications identified overlapping target autoantigens or further validated the identified target autoantigens by orthogonal techniques to eliminate potential false positives that are known occur with autoantigen arrays [57]. In one study, serum samples taken exclusively from MIS-C subjects before they received IVIG identified several autoantibodies including high levels against three members of the casein kinase family (CSNK1A, CSNK2A1, and CSNK1E) [18]. However, no validation immunoassays were used to independently confirm the relevance of these targets and the number of negative controls was very limited. Using the sensitive and specific LIPS technology, we were unable to detect any CSNK1-A autoantibodies, the previous study’s most informative biomarker, in any of our MIS-C patients not receiving IVIG (Burbelo, unpublished), raising questions about the usefulness or validity of these autoantibodies as markers of MIS-C. In the case of the two additional antigen array publications discovering autoantibodies in MIS-C, one study identified seropositive La, Jo-1, and IL-1α autoantibodies in their MIS-C cohort [17] and the other study identified gastric ATPase and Ro60 (TROVE2) as the two most abundant autoantibodies in patients with MIS-C [22]. Our findings showing the lack of La, Ro60 and gastric ATPase autoantibodies in children with MIS-C who did not receive IVIG, but high levels of these autoantibodies in MIS-C patients receiving IVIG, suggests that detection of several autoantibodies in the two previously published studies might have been due to the IVIG given to the patients [17, 22]. Future autoantibody studies utilizing larger cohorts of disease controls and MIS-C patients before IVIG administration coupled with follow-up independent immunoassay validation are needed [58].

The exact mechanisms for the therapeutic benefit of IVIG in autoinflammatory conditions likely involves multiple effects including the presence of anti-pathogen neutralizing antibodies, action on Fc receptors, and effects on complement and other immunomodulatory effects [13, 59]. Most recently, IVIG has been shown to mediate neutrophil cell death through a Fab2-mediated mechanism [59]. Although we describe at least four autoantibodies with relatively high levels present in IVIG preparations that can be potentially passively transferred to patients with MIS-C, KD, and other diseases, their functional activity, if any, i*n vivo* remains unknown. Based on the robust detection of the IVIG-associated autoantibodies, the *in vivo* decay of these autoantibodies was studied. From our analysis of MIS-C IVIG recipients, Ro52, Ro60, and La autoantibodies showed a similar decay profile, in which all three autoantibodies were present in the blood for several weeks before becoming undetectable by 35-60 days. In contrast, the decay kinetics of autoantibodies directed against the gastric ATPase was much longer (>100 days). One potential explanation for this decay discrepancy is that Ro52, Ro60, and La are all intracellular proteins [60], and the shorter autoantibody decay may reflect limited persistence in specific tissue due to inaccessibility of the antibodies to the intracellular compartment where these antigens are located. In contrast, gastric ATPase is a surface membrane protein that is antibody accessible, which might allow gastric ATPase autoantibodies to bind gastric tissue and thereby slowly be released resulting in persistent presence in the blood. Based on the availability of serial samples from IVIG recipients and additional studies of the decay of both anti-pathogen and autoantibodies present in preparations, further insights may be gained into the therapeutic action of IVIG.

In conclusion, unlike adults with COVID-19, our study found no evidence that children with MIS-C or COVID-19 produce autoantibodies associated with known autoimmune diseases. Since we only examined a limited panel of autoantigens, we appreciate that there may be other autoantibodies with MIS-C that we did not detect. In light of recent studies showing increased somatic hypermutation in plasmablasts of MIS-C [23, 61], additional studies are needed in MIS-C to determine if specific autoantibodies, play a role in immune complex formation or autoantibody-driven damage to host cells.

## Data Availability

All data produced in the present work are contained in the manuscript.

## Acknowledgements

This work was supported in part by the Intramural Research Programs of NIDCR and NIAID of the National Institutes of Health and Regione Lombardia (project “Immune response in patients with COVID-19 and co-morbidities”), FONDECYT 11181222 (M.C.P.), ANID-COVID19 0999 (M.C.V.) and NIH 3R01HL140898-03S1 and NICHD 1R61HD105590 (J.C.B and A.H.T). We thank the NIAID Office of Cyber Infrastructure and Computational Biology, Bioinformatics and Computational Biosciences Branch (Contract HHSN316201300006W/HHSN27200002 to MSC, Inc) and Operations Engineering Branch for developing the HGRepo system to enable streamlined access to the data and the NCI Advanced Biomedical Computational Science (ABCS) for data transformation support. The content of this publication does not necessarily reflect the views or policies of the Department of Health and Human Services, nor does mention of trade names, commercial products, or organizations imply endorsement by the U.S. Government.

